# Awareness, digital literacy and institutional factors associated with self-reported utilization of the UpToDate clinical decision support tool among lecturers in Kenya Medical Training College

**DOI:** 10.64898/2026.11.20.26368626

**Authors:** Kainde Musyoka, Ben Kinyua, Regina Kombo, George Tibi, George Adie

## Abstract

**Background:** Digital clinical decision support tools such as UpToDate can strengthen evidence-based teaching and practice, but access does not guarantee use. UpToDate has been available at selected Kenya Medical Training College (KMTC) campuses since 2018 yet recorded use is very low. We examined factors associated with UpToDate utilization among KMTC lecturers.

**Methods:** We conducted a cross-sectional survey in August to September 2026 among 215 lecturers at campuses with institutional UpToDate access. A self-administered questionnaire was used to measure awareness, digital literacy, self-efficacy, institutional factors and a five-item self-rated utilization index. We used t-tests, ANOVA, Pearson correlation and hierarchical linear regression with HC3 robust standard errors, and content analysis.

**Results:** Although 79.1% had heard of UpToDate and 70.7% knew KMTC provides access, only 5.6% rated their awareness as high. UpToDate-specific digital literacy was low, moderate and high in 10.2%, 60.0% and 29.8%. Mean utilization was lower among females (2.91 vs 3.17) and declined with age (3.21 at 23 to 34 years to 2.05 at more than 55 years). Digital literacy (r = 0.49) and self-efficacy (r = 0.44) correlated positively with utilization. Institutional factors correlated with utilization (r = 0.37 to 0.49). In the fully adjusted model, older age more than 55 years (b = −1.33, 95% CI −1.96 to −0.70), more than 16 years teaching experience (b = 0.62) and principal-lecturer rank (b = 0.61) remained independently associated with utilization.

**Conclusion:** There is access to UpToDate at KMTC but the conversion of access into routine use is a challenge. Sustained awareness-building, hands-on training, departmental champions and integration into teaching and supervision are priorities to enhance utilization.

## INTRODUCTION

Evidence-based practice depends on clinicians and educators being able to find, appraise and apply current evidence quickly. Point-of-care clinical decision support tools such as UpToDate, which provide regularly updated, peer-reviewed topic reviews and drug information, are intended to make that possible (Wolters Kluwer, 2026). Use of UpToDate has been associated with better hospital outcomes in the United States, and the resource is available in more than 190 countries (Isaac et al., 2012; Kinengyere et al., 2021). In low- and middle-income countries (LMICs), where textbooks and journal subscriptions are often out of date or unaffordable, donated and discounted institutional licenses, notably through the Better Evidence Program, have extended access to health professions institutions across sub-Saharan Africa (Kinengyere et al., 2021).

Access, however, is not use. In Uganda, UpToDate recorded 43,043 log-ins and 15,591 registrations at Makerere University College of Health Sciences in its first year of free access. However, the authors described uptake as inconsistent and use as low relative to the eligible population (Kinengyere et al., 2021). A study in Tanzania found moderate awareness, insufficient knowledge and search skills, and barriers including a cumbersome registration process, poor internet quality and lack of time (Adam & MKHAI, 2025a). In a cohort of 1,681 clinicians in limited-resource settings who were given free UpToDate subscriptions, difficulty fitting the tool into the workflow was the only barrier significantly associated with lower use, while a supportive professional context and perceived usefulness were associated with greater use (Rosenberg et al., 2022). A recent meta-analysis of health-worker adoption of digital health technologies in LMICs similarly identified facilitating conditions as the strongest correlate of actual use (Wang et al., 2025a). Evidence on Kenyan health professions educators is thin. Related Kenyan work has addressed evidence-based medicine practice among resident physicians and, at KMTC, e-learning and faculty digital equity, but not use of a clinical decision support tool. (Gachanja et al., 2021; Gicheru & Mwangi, 2023; Unadkat et al., 2021a)

Lecturers are a strategically important group. They teach, supervise students in clinical placements and model information-seeking behavior. If they do not use evidence tools, students are unlikely to adopt them. Unlike clinicians, whose use is shaped by patient flow and point-of-care time pressure (Rosenberg et al., 2022), lecturers’ use should depend more on awareness, skills, departmental norms and curricular integration. Therefore an understanding of awareness and actual utilization among lecturers in a middle-level health training institution is central to enhancing uptake.

KMTC is the largest public provider of middle-level health professions training in Kenya and is reported to train most of the country’s middle-level health workforce (KMTC, 2023). Selected campuses gained institutional UpToDate access between 2018 and 2021. Campus librarians were trained to cascade its use, and digital transformation is a stated priority of the 2023–2028 strategic plan (KMTC, 2023). However, there is little evidence on why an already-funded resource is so little used and which levers are likely to change this. Therefore, we describe awareness of, and UpToDate-specific digital literacy among KMTC lecturers, as well as identify individual characteristics and institutional factors associated with utilization.

## METHODS

### Study design and reporting

We conducted a descriptive, analytical cross-sectional survey with an embedded qualitative component (a single open-ended item, analysed concurrently and used to contextualize the quantitative findings) (Creswell JW & Plano Clark VL., 2018). Data was collected from August to September 2026. The study is reported according to the STROBE guideline for cross-sectional studies.

### Setting

The study took place at 22 KMTC campuses that have institutional UpToDate (Wolters Kluwer) access. Access was introduced between 2018 and 2021 through discounted licenses arranged with the Better Evidence Program, and campus librarians were trained to promote it to staff and students. At the time of the study UpToDate was the only clinical decision support tool in routine institutional use at KMTC and was accessed through campus libraries and institutional networks.

### Participants and sampling

The target population was lecturers teaching in any KMTC department at campuses with UpToDate access, including full-time KMTC employees and lecturers seconded by county governments (sampling frame N = 654). Lecturers were eligible if they had taught at the campus for more than one semester and gave written informed consent. Part-time lecturers and those on sick, maternity or paternity leave or away on official duty throughout data collection were excluded. Participants were drawn by simple random sampling from the campus staff lists using random numbers, with the sample drawn proportionally across campuses. Ineligible or unreachable lecturers were replaced by the next randomly selected lecturer. Lecturers who took part in the questionnaire pretest survey were removed from the frame.

The required sample was calculated with Cochran’s formula for estimating a proportion in a single population.

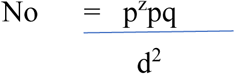

Where Z=1.96(95% confidence level)

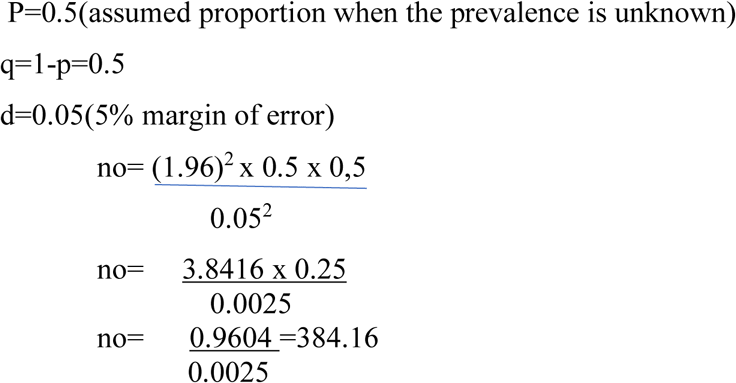

Since the total population is less than10,000 adjustment of the sample size is done using

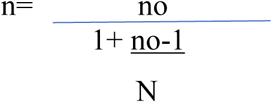

Where no=384 N=654

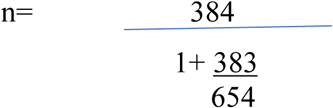

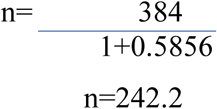

The sample size will consist of 242 lecturers.

To cater for non -response, the final sample size was adjusted by 10% (242 +24=**266)** Therefore, the final sample size was 266 participants

### Instrument and variables

A structured, self-administered English questionnaire had five sections: (i) demographic and professional characteristics (gender, age group, department, job title, administrative role, years of teaching experience, prior formal UpToDate training); (ii) awareness of UpToDate (ever heard of it; knowledge that KMTC provides access; self-rated prior awareness); (iii) digital literacy in relation to UpToDate (familiarity with its functions and understanding of how to use it effectively); (iv) individual characteristics (attitude toward technology, self-efficacy/perceived capability); and (v) institutional factors (reliable internet access, reliable work-device access, departmental encouragement, integration into teaching and clinical follow-up, on-site training, institutional support, clear policies or guidelines, and leadership support). The questionnaire ended with one open-ended item inviting additional comments. Digital literacy scores were classified as Low (1.0–2.0), Moderate (2.5–3.5) or High (4.0–5.0).

Attitude, self-efficacy, digital literacy, institutional and utilization items used five-point Likert responses (1 = strongly disagree to 5 = strongly agree). Composite scores were the mean of their items (range 1–5; higher = more of the construct). The primary outcome was the utilization index, the mean of five Likert items on self-rated functional use of UpToDate: ability to access the platform on a computer or smartphone, to log in with an institutional account, to search efficiently, to interpret and apply retrieved information in teaching or clinical practice, and to navigate its sections unaided. The index therefore captures self-reported practical use competence and not the frequency or duration of use. Self-efficacy comprised confidence in evaluating retrieved information, perceived adequacy of training and perceived digital competence. Digital literacy comprised familiarity with UpToDate’s functions and understanding of how to use it effectively. Internal consistency was assessed with Cronbach’s α, with 0.70 taken as acceptable (Table 1).

**Table 1.** Composite scales and internal-consistency reliability.

| **Scale** | **k items** | **Cronbach's α** | **Role in analysis** |
|---|---|---|---|
| Attitude toward technology/UpToDate | 8 | 0.817 | Individual characteristic |
| Digital literacy regarding UpToDate | 2 | 0.687 | Awareness/literacy (Obj. 1) |
| Self-efficacy / perceived capability | 3 | 0.723 | Individual characteristic (Obj. 2) |
| Utilization Index (self-rated) | 5 | 0.842 | Dependent variable |
| Institutional factors composite | 8 | 0.816 | Institutional factor (Obj. 3) |

### Pre-testing, data collection and management

Face and content validity were assessed by two public health researchers who judged the relevance, clarity and coverage of each item. The questionnaire was pre-tested among about 10% of the sample size, among lecturers who were not part of the main sample, to check clarity, completion time and item order. The instrument was revised and pretest data were excluded. Data collectors administered paper questionnaires and entered responses into Microsoft Excel with double verification. The dataset was checked for duplicates, out-of-range, inconsistent and implausible values against the original forms, and exported to Stata 14 (StataCorp, College Station, TX). A codebook was used to define all variables and reference categories.

### Statistical analysis

Categorical variables are summarised as frequencies and percentages and composite scores as means and standard deviations. Mean utilization was compared across groups with independent-samples t-tests (two categories) and one-way ANOVA (more than two categories). Effect sizes were calculated from the reported test statistics as Cohen’s d (t-tests) and η² (ANOVA). Pearson’s r with 95% confidence intervals (Fisher z transformation) quantified associations between utilization and continuous predictors.

To estimate independent associations, we fitted hierarchical ordinary least squares regression models with the utilization index as outcome. Model 1 included gender, age group, teaching experience, job title, prior formal UpToDate training, attitude toward technology, digital literacy and self-efficacy (13 coefficients). Model 2 added the institutional factors composite. We used heteroskedasticity-consistent (HC3) standard errors [20] (Stata: *regress, vce(hc3)*) and report unstandardised coefficients (b), robust standard errors, 95% confidence intervals, R², root mean squared error and the overall F-test. Department was not entered in the models. Tests were two-sided with α = 0.05. Analyses were not adjusted for multiple comparisons and should be regarded as exploratory. Multicollinearity was checked with variance inflation factors (all < 4). Complete-case analysis was used. Because two institutional items (reliable internet and device access; both scored 1 = strongly disagree to 5 = strongly agree) were unexpectedly inversely associated with utilization, we examined inter-item correlations, compared lecturers who strongly agreed with either item with the rest, examined the association within blocks of the questionnaire serial number (which follows the order of data collection by campus) and repeated Model 2 using only the six departmental, training, policy and leadership items. Open-ended responses were analysed by conventional (inductive) content analysis. Comments were read repeatedly, open codes were generated manually and grouped into categories and then themes.

### Ethical considerations

The study was approved by the KMTC Institutional Research and Ethics Review Committee and the Chuka University Ethics Committee for Research, and a research permit was issued by the National Commission for Science, Technology and Innovation (NACOSTI). Campus principals authorized data collection. All participants received an information sheet and gave written informed consent. Participation was voluntary, without incentives, and questionnaires carried only a serial number; no names or staff numbers were recorded. Forms and files were stored securely and results are reported in aggregate.

## RESULTS

### Respondent Characteristics

Of 266 lecturers sampled, 215 completed the questionnaire (response rate 80.8%). Slightly over half (118, 54.9%) of respondents were male. Most were aged 23–34 years (86, 40.6%), drawn mostly from nursing (81, 37.7%) and public health (46, 21.4%) departments. Majority (164, 76.3%) were holding the rank of lecturer, and nearly half (98, 45.6%) had 6–10 years’ teaching experience. This is as summarized in table 2

**Table 2:**
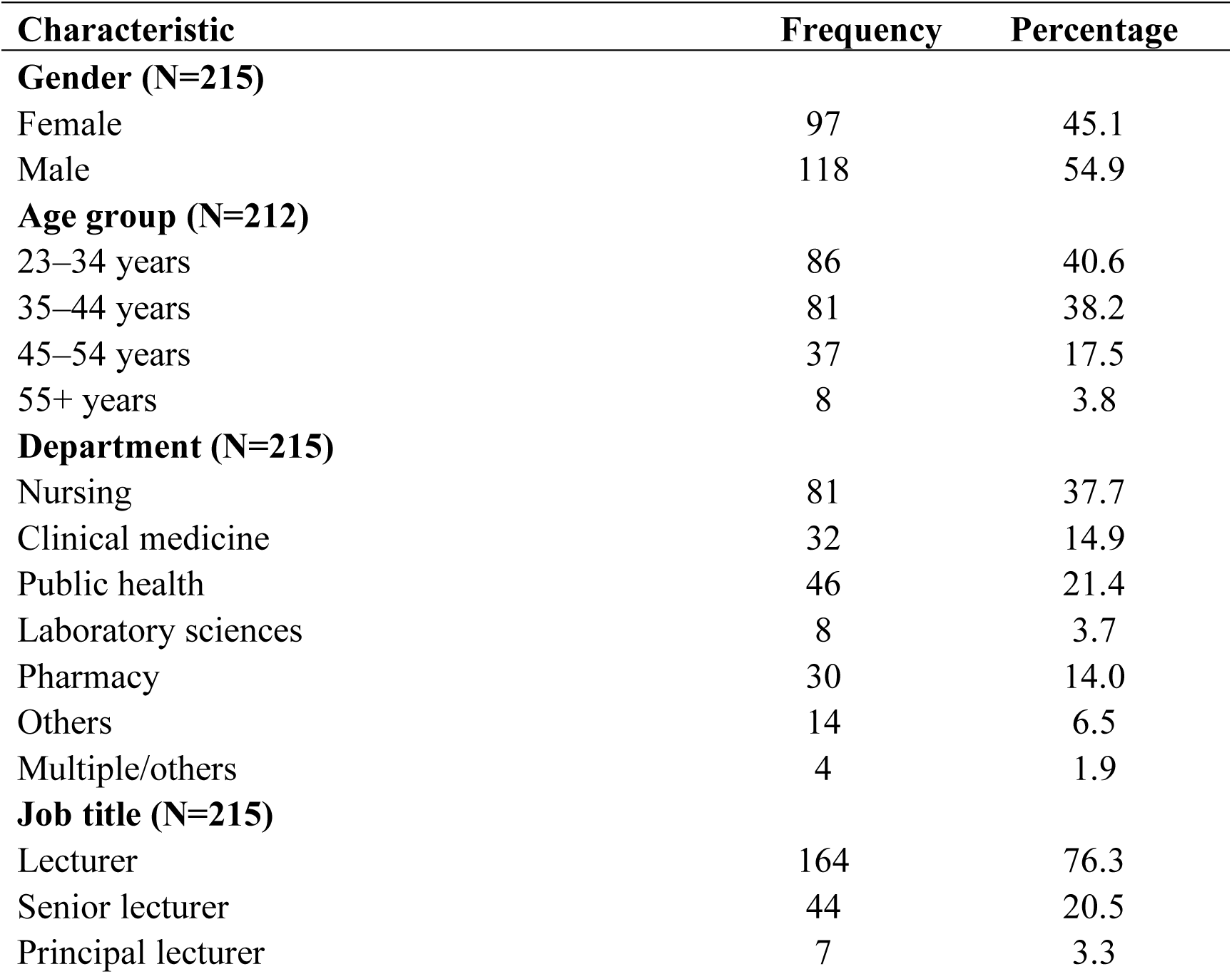
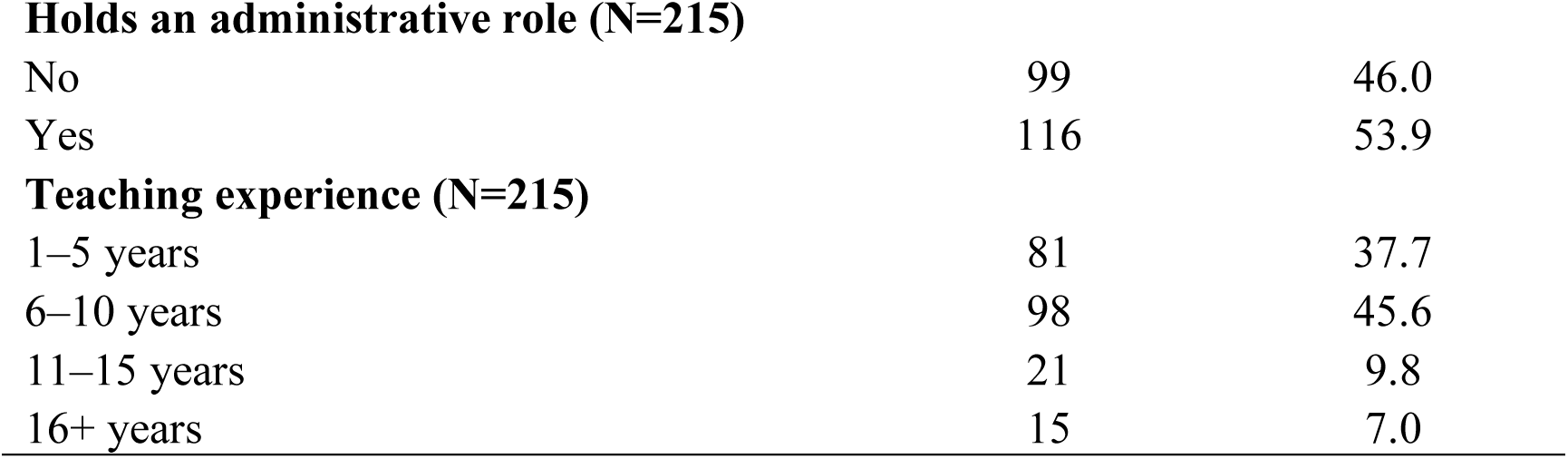
Respondent Characteristics (N=215)

| Characteristic | Frequency | Percentage |
|---|---|---|
| **Gender (N=215)** |  |  |
| Female | 97 | 45.1 |
| Male | 118 | 54.9 |
| **Age group (N=212)** |  |  |
| 23–34 years | 86 | 40.6 |
| 35–44 years | 81 | 38.2 |
| 45–54 years | 37 | 17.5 |
| 55+ years | 8 | 3.8 |
| **Department (N=215)** |  |  |
| Nursing | 81 | 37.7 |
| Clinical medicine | 32 | 14.9 |
| Public health | 46 | 21.4 |
| Laboratory sciences | 8 | 3.7 |
| Pharmacy | 30 | 14.0 |
| Others | 14 | 6.5 |
| Multiple/others | 4 | 1.9 |
| **Job title (N=215)** |  |  |
| Lecturer | 164 | 76.3 |
| Senior lecturer | 44 | 20.5 |
| Principal lecturer | 7 | 3.3 |
| **Holds an administrative role (N=215)** |  |  |
| No | 99 | 46.0 |
| Yes | 116 | 53.9 |
| **Teaching experience (N=215)** |  |  |
| 1–5 years | 81 | 37.7 |
| 6–10 years | 98 | 45.6 |
| 11–15 years | 21 | 9.8 |
| 16+ years | 15 | 7.0 |

### Awareness and digital literacy of Kenya Medical Training College lecturers on Up-to-Date clinical decision resource tool

Majority of respondents (170, 79.1%) had heard of the UpToDate and aware that that KMTC provides institutional access (152, 70.7%). However, only 5.6% rated their own awareness prior to this study as “High”. (Figure 1)

**Figure 1:**
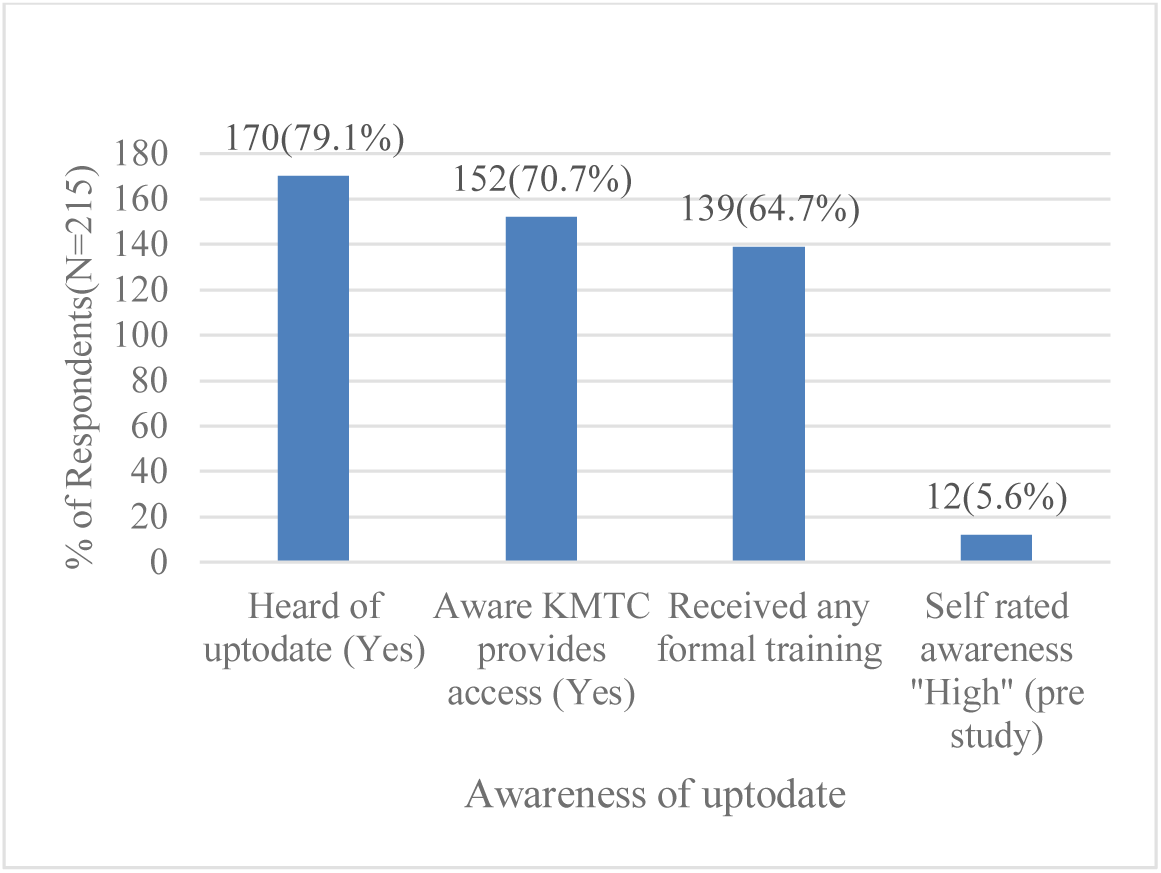
**Awareness of up-to-date clinical decision support tool**

The digital-literacy composite (familiarity with UpToDate’s functions and understanding of how to use it effectively; M = 3.32, SD = 0.64, range 1–5), was classified as high by a third of lecturers (64, 29.8%), moderate in 129(60.0%), and low in 22(10.2%). This indicates that digital literacy specific to using it effectively lags behind, with the majority of lecturers only moderately literate (Figure 2).

**Figure 2:**
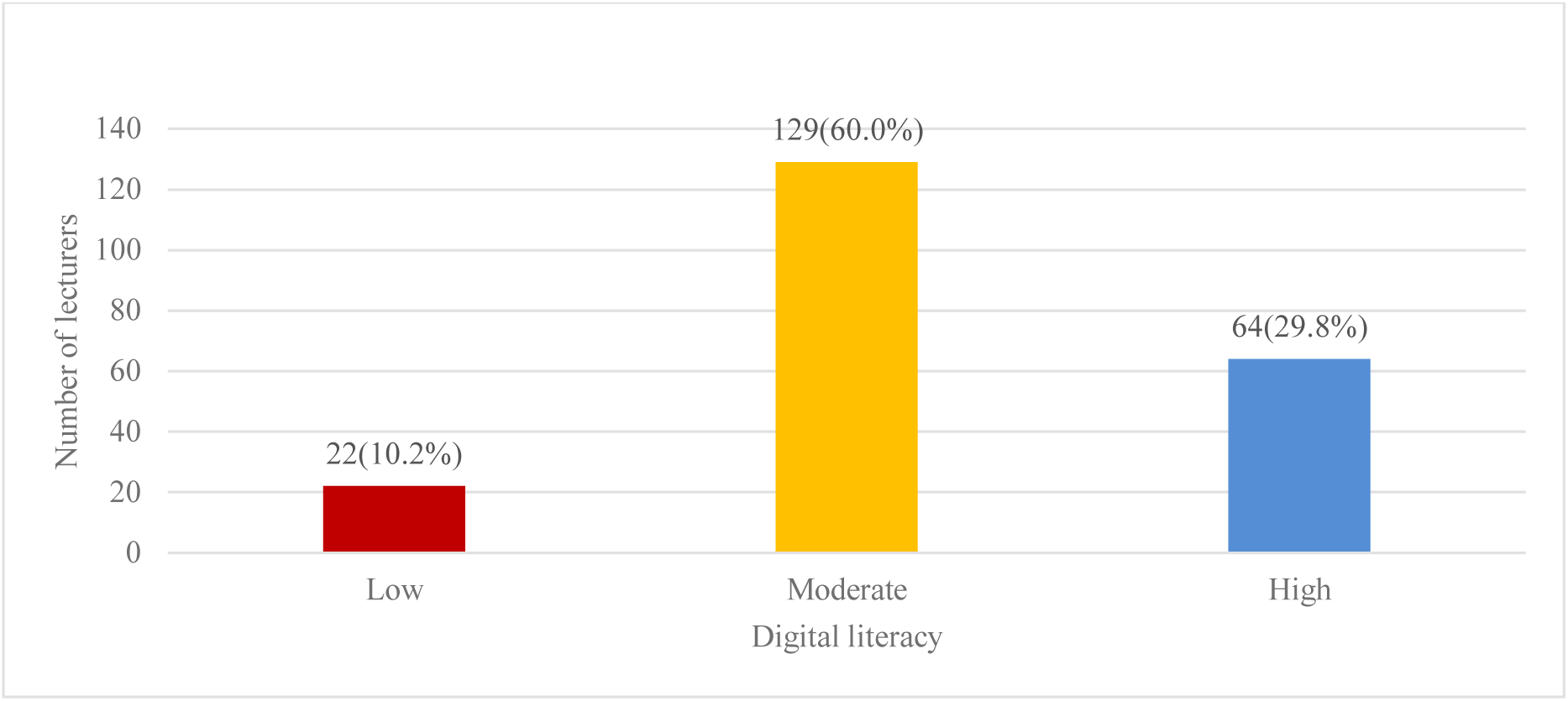
**Digital literacy regarding up-to-date (composite of familiarity &understanding items)**

**Figure 3:**
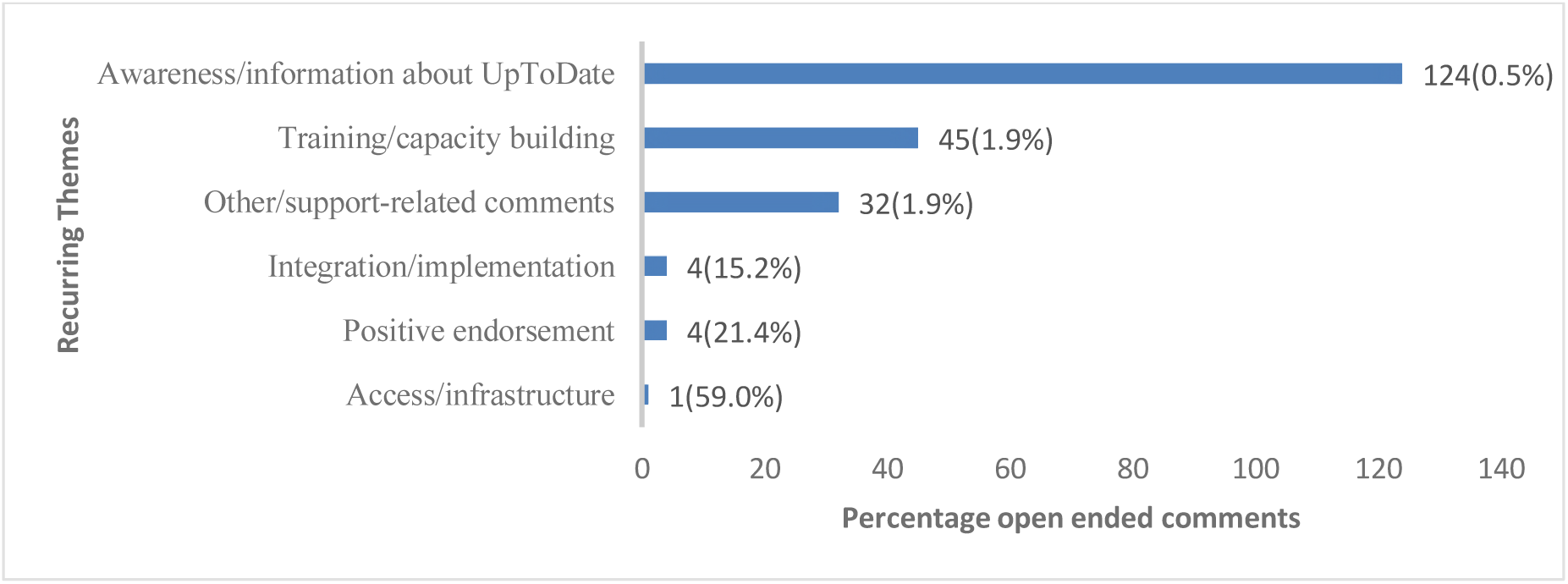
**Recurring themes in open ended comments on up-to-date clinical support tool**

### Individual characteristics and the utilization of Up-to -Date clinical decision resource tool among lecturers at the Kenya Medical Training College

Independent-samples t-tests were used for two-category variables and one-way ANOVA for variables with more than two categories. Utilization differed significantly by gender. Men scored higher than women (3.17 vs 2.91; t = −2.78, p = 0.006). Utilization fell with age, from 3.21 among those aged 23–34 years to 2.05 among those aged ≥55 (F = 7.92, p < 0.001), and rose with rank from lecturer (2.99) to senior lecturer (3.20) and principal lecturer (3.54) (F = 3.67, p = 0.027). Lecturers with prior formal training scored markedly higher than those without (3.29 vs 2.60; t = 7.98, p < 0.001). We found no significant differences by administrative role (p = 0.364), teaching experience (p = 0.310) or department (p = 0.054), although the departmental difference was of small-to-moderate magnitude. This is summarized in table 3

**Table 3.** Utilization index of the UpToDate by individual characteristics.

| Variable / group | n | M (Utilization) | SD | Test statistic | p-value |
|---|---|---|---|---|---|
| **Gender** |  |  |  | t = −2.779 | 0.006 |
| Female | 97 | 2.91 | 0.72 |  |  |
| Male | 118 | 3.17 | 0.65 |  |  |
| **Age group (years)** |  |  |  | F = 7.92 | <0.001 |
| 23–34 | 86 | 3.21 | 0.56 |  |  |
| 35–44 | 81 | 3.01 | 0.74 |  |  |
| 45–54 | 37 | 2.98 | 0.62 |  |  |
| 55+ | 8 | 2.05 | 1.01 |  |  |
| **Department** |  |  |  | F = 2.11 | 0.054 |
| Nursing | 81 | 2.95 | 0.78 |  |  |
| Clinical Medicine | 32 | 3.09 | 0.79 |  |  |
| Public Health | 46 | 3.07 | 0.44 |  |  |
| Laboratory Sciences | 8 | 2.55 | 0.83 |  |  |
| Pharmacy | 30 | 3.2 | 0.49 |  |  |
| Others | 14 | 3.29 | 0.75 |  |  |
| Multiple/Other | 4 | 3.65 | 0.41 |  |  |
| **Job title** |  |  |  | F = 3.67 | 0.027 |
| Lecturer | 164 | 2.99 | 0.67 |  |  |
| Senior Lecturer | 44 | 3.2 | 0.75 |  |  |
| Principal Lecturer | 7 | 3.54 | 0.44 |  |  |
| **Administrative role** |  |  |  | t = −0.909 | 0.364 |
| No | 99 | 3 | 0.73 |  |  |
| Yes | 116 | 3.09 | 0.66 |  |  |
| **Prior formal UpToDate training** |  |  |  | t = −7.975 | <0.001 |
| No | 76 | 2.60 | 0.75 |  |  |
| Yes | 139 | 3.29 | 0.52 |  |  |
| **Teaching experience (years)** |  |  |  | F = 1.20 | 0.31 |
| 1–5 | 81 | 3.11 | 0.66 |  |  |
| 6–10 | 98 | 3 | 0.64 |  |  |
| 11–15 | 21 | 2.9 | 0.81 |  |  |
| 16+ | 15 | 3.25 | 0.96 |  |  |

Pearson correlation coefficients were computed to examine the strength and direction of the linear association between each continuous predictor and the utilization index (Table 4). Utilization correlated positively with digital literacy (r = 0.487, p < 0.001) and self-efficacy /capability (r = 0.445, p < 0.001) but not with attitude toward technology (r = −0.038, p =0.577).

**Table 4.** Pearson correlations with Utilization Index.

| **Predictor** | **Pearson's r** | **p-value** |
|---|---|---|
| Digital literacy | 0.487 | <0.001 |
| Attitude toward technology | −0.038 | 0.577 |
| Self-efficacy/capability | 0.445 | <0.001 |

### Institutional factors and utilization of Up-to-Date clinical decision resource tool among lecturers at the Kenya Medical Training College

All eight institutional items were significantly correlated with utilization (Table 5). Positive associations were observed for departmental meetings/encouragement (r = 0.481), integration of UpToDate into teaching and clinical follow-up (r = 0.477), campus institutional support (r = 0.485), on-site training (r = 0.444), clear policies and guidelines (r = 0.443) and leadership support (r = 0.371). The institutional composite correlated with utilization at r = 0.456 (0.34–0.56). In contrast, reliable institutional internet access (r = −0.268) and reliable work-device access (r = −0.284) were inversely correlated with utilization.

**Table 5:** Pearson correlation between institutional factors and uptodate utilization.

| **Institutional Item** | **Pearson's r** | **95% CI** | **p-value** |
|---|---|---|---|
| Reliable institutional internet access | −0.268 | −0.39 to −0.14 | <0.001 |
| Reliable work-device access | −0.284 | −0.40 to −0.16 | <0.001 |
| Departmental meetings/encouragement on UpToDate | 0.481 | 0.37 to 0.58 | <0.001 |
| Integration of UpToDate into teaching/clinical follow-up | 0.477 | 0.37 to 0.57 | <0.001 |
| On-site campus training | 0.444 | 0.33 to 0.55 | <0.001 |
| Campus/institutional support for lecturers | 0.485 | 0.38 to 0.58 | <0.001 |
| Clear institutional policies/guidelines | 0.443 | 0.33 to 0.54 | <0.001 |
| Leadership/administrative support | 0.371 | 0.25 to 0.48 | <0.001 |
| Institutional factors composite (8 items) | 0.456 | 0.34 to 0.56 | <0.001 |

**Table 6:** Pearson correlation between individual factors and UpToDate utilization.

| **Predictor** | **Pearson's r** | **95% CI** | **p** |
|---|---|---|---|
| Digital literacy (UpToDate-specific) | 0.487 | 0.38 to 0.58 | <0.001 |
| Self-efficacy / perceived capability | 0.444 | 0.33 to 0.55 | <0.001 |
| Attitude toward technology | −0.045 | −0.18 to 0.09 | 0.516 |

**Table 6.** Hierarchical linear regression of the utilization index (HC3 robust standard errors)

| Predictor | Model 1 b (SE) | 95% CI | p-value | Model 2 b (SE) | 95% CI | p-value |
|---|---|---|---|---|---|---|
| Intercept | 2.604 (0.538) | [1.54, 3.67] | **<0.001** | 2.400 (0.696) | [1.03, 3.77] | **0.001** |
| Gender |  |  |  |  |  |  |
| Female (ref: Male) | -0.158 (0.082) | [-0.32, 0.00] | 0.055 | -0.161 (0.082) | [-0.32, 0.00] | **0.051** |
| Age group |  |  |  |  |  |  |
| 35–44 years (ref: 23–34) | -0.237 (0.110) | [-0.45, -0.02] | **0.033** | -0.234 (0.109) | [-0.45, -0.02] | **0.034** |
| 45–54 years (ref: 23–34) | -0.300 (0.125) | [-0.55, -0.05] | **0.017** | -0.311 (0.128) | [-0.56, -0.06] | **0.016** |
| 55+ years (ref: 23–34) | -1.324 (0.313) | [-1.94, -0.71] | **<0.001** | -1.342 (0.305) | [-1.94, -0.74] | **<0.001** |
| Teaching experience |  |  |  |  |  |  |
| 6–10 years (ref: 1–5) | 0.040 (0.111) | [-0.18, 0.26] | 0.721 | 0.046 (0.113) | [-0.18, 0.27] | 0.682 |
| 11–15 years (ref: 1–5) | -0.049 (0.164) | [-0.37, 0.27] | 0.765 | -0.046 (0.166) | [-0.37, 0.28] | 0.78 |
| 16+ years (ref: 1–5) | 0.646 (0.189) | [0.27, 1.02] | **0.001** | 0.630 (0.188) | [0.26, 1.00] | **0.001** |
| Job title |  |  |  |  |  |  |
| Senior Lecturer (ref: Lecturer) | 0.219 (0.121) | [-0.02, 0.46] | 0.071 | 0.234 (0.126) | [-0.01, 0.48] | 0.065 |
| Principal Lecturer (ref: Lecturer) | 0.616 (0.277) | [0.07, 1.16] | **0.027** | 0.624 (0.278) | [0.08, 1.17] | **0.026** |
| UpToDate training |  |  |  |  |  |  |
| No (ref: Yes) | -0.264 (0.135) | [-0.53, 0.00] | 0.052 | -0.188 (0.164) | [-0.51, 0.14] | 0.252 |
| Individual-level factors |  |  |  |  |  |  |
| Attitude toward technology | -0.028 (0.120) | [-0.27, 0.21] | 0.816 | -0.034 (0.118) | [-0.27, 0.20] | 0.774 |
| Digital literacy | 0.141 (0.091) | [-0.04, 0.32] | 0.124 | 0.130 (0.094) | [-0.05, 0.31] | 0.168 |
| Self-efficacy/capability | 0.148 (0.093) | [-0.04, 0.33] | 0.115 | 0.124 (0.087) | [-0.05, 0.30] | 0.153 |
| Institutional-level factors |  |  |  |  |  |  |
| Institutional support | — | — | — | 0.091 (0.124) | [-0.15, 0.33] | 0.464 |

Pearson correlation analysis showed statistically significant positive associations between UpToDate utilization and digital literacy (r = 0.487, p < 0.001), institutional factors (r = 0.453, p < 0.001), and self-efficacy/capability (r = 0.445, p < 0.001). In contrast, attitude toward technology was not significantly associated with UpToDate utilization (r = −0.038, p = 0.577).

### Hierarchical regression analysis of factors associated with UpToDate utilization

A hierarchical linear regression analysis was conducted to examine the independent associations between individual characteristics, individual-level factors, and institutional support and utilization of the UpToDate clinical decision support tool. The utilization index was entered as the continuous dependent variable (N = 212). Model 1 included demographic and individual-level predictors including gender, age, teaching experience, job title, formal UpToDate training, attitude toward technology, digital literacy, and self-efficacy/capability. Model 2 additionally included the institutional factors composite. Heteroskedasticity-consistent HC3 standard errors were used to provide robust inference.

Model 1 (n = 212) explained 41.1% of the variance in utilization (R² = 0.411; F(13,198) = 14.06, *p* < 0.001; root MSE = 0.550; Table 4.5). Compared with lecturers aged 23–34 years, utilization was lower among those aged 35–44 years (b = −0.237, *p* = 0.033), 45–54 years (b = −0.300, *p* = 0.017) and ≥55 years (b = −1.324, *p* < 0.001). Lecturers with ≥16 years of teaching experience scored higher than those with 1–5 years (b = 0.646, *p* = 0.001), as did principal lecturers relative to lecturers (b = 0.616, *p* = 0.027). Female gender (b = −0.158, *p* = 0.055), no prior formal UpToDate training (b = −0.264, *p* = 0.052), attitude, digital literacy and self-efficacy were not significantly associated with utilization.

Adding the institutional support composite (Model 2) changed R² only from 0.411 to 0.414 (F(14,197) = 14.87, *p* < 0.001). The composite was not independently associated with utilization (b = 0.091, 95% CI −0.15 to 0.33, *p* = 0.464), and the associations of age (≥55 years: b = −1.342, *p* < 0.001), ≥16 years of teaching experience (b = 0.630, *p* = 0.001) and principal-lecturer rank (b = 0.624, *p* = 0.026) were essentially unchanged. The coefficient for no prior training weakened (b = −0.188, p = 0.252), and was no longer statistically significant.

### Supplementary Content Analysis of Open-Ended Comments

A brief content analysis of the substantive comments from respondents identified several recurring themes. The most common theme was awareness and information about the UpToDate clinical decision support tool, reported by over half (124, 59.0%). Respondents frequently requested greater awareness, sensitization, introduction to the tool, and more information about its functionality and use. Typical comments included requests to “create awareness,” “get more information about the tool,” and understand “what the tool entails.”

The second major theme was training and capacity building (45, 21.4%). Respondents requested training on how to use UpToDate, including onsite or physical training, improved learning sessions, mentorship, and regular training opportunities. A smaller number of comments related to other support or implementation issues (32, 15.2%), including requests for continued support, updates, improved teaching or teamwork, and related suggestions. Positive endorsement of the tool was expressed in four comments (1.9%), with respondents describing UpToDate as effective, useful, or a good initiative. Integration and implementation issues were mentioned in four comments (1.9%), including suggestions concerning LMS integration, learner access, and broader institutional adoption. Access or infrastructure-related issues were identified in one comment (0.5%). The findings complement the quantitative results by suggesting that limited awareness and inadequate training may represent important barriers to effective utilization of the clinical decision support tool. (Figure 4.3)

## DISCUSSION

### Overview

This is, to our knowledge, the first study of utilization of a clinical decision support tool among lecturers in a Kenyan middle-level health training institution. Four findings stand out. First, nominal awareness was high but confident familiarity was rare. Second, attitude toward technology was unrelated to use, whereas capability (digital literacy, self-efficacy) was positively correlated. Third, age, rank and long teaching experience were independently associated with use in adjusted models. Fourth, all six support-related institutional items (departmental encouragement, integration into teaching, training, support, policy and leadership) were positively correlated with use.

### Awareness and digital literacy relating to the UpToDate clinical decision support tool

The findings revealing that 79.1% of lecturers had heard of UpToDate but only 5.6% rated their prior awareness as high suggests that depth of awareness at KMTC is low. Lecturers know the resource exists but have limited knowledge on what it contains or how to use it. This echoes Uganda, where uptake remained inconsistent despite free access and librarian-led promotion (Kinengyere et al., 2021). Similarly, in Tanzania, awareness was moderate and knowledge and search skills were insufficient to generate strong uptake, particularly where users were unfamiliar with the resource(Adam & MKHAI, 2025b). The finding is important because a clinical decision support tool can only be useful if users know how to access it, search for relevant information and apply the information obtained to their teaching and clinical activities.

Data from KMTC faculty indicate substantial digital access (75.9% had internet at work and 75.2% a work laptop) but only a minority had advanced skills in the learning management system (Gicheru & Mwangi, 2023). Our finding that most lecturers were only moderately literate in UpToDate-specific use is consistent with that gap between access and advanced competence. The open-ended comments, in which awareness/information (59.0%) and training (21.4%) dominated, came from respondents themselves and strengthens the interpretation of the quantitative findings. It is probable that the availability of institutional access therefore does not necessarily mean that all potential users are equally prepared to make full use of the resource. The present findings therefore suggest that providing institutional access may need to be accompanied by practical guidance on how to access, search and use UpToDate effectively.

A previous KMTC-focused study by Gachanja (2021), identified challenges relating to ICT proficiency, infrastructure and access to digital teaching materials during the transition to digital education (Gachanja et al., 2021). Similarly, Gicheru and Mwangi (2023) emphasized the need to institutionalize digital equity within KMTC so that differences in access and confidence do not limit the benefits of digital technologies (Gicheru & Mwangi, 2023). Our study revealed that digital literacy was moderately and positively correlated with UpToDate utilization (r=0.487, p<0.001). Self-efficacy showed a similar positive relationship with utilization (r=0.445, p<0.001).

The hierarchical regression, however, indicated that after adjustment for demographic and other individual characteristics, digital literacy and self-efficacy were no longer statistically significant independent predictors of utilization. This does not necessarily mean that digital literacy and self-efficacy are unimportant. Rather, it suggests that their relationship with utilization may overlap with other characteristics such as age, teaching experience, job position and training. It is therefore possible that digital competence and confidence operate together with other individual and institutional factors. Overall, the findings suggest that there is a gap between knowing about UpToDate and being fully prepared to use it. Therefore, continued awareness creation, practical training and capacity building may be important for increasing effective use of UpToDate at KMTC.

### Individual characteristics contributing to UpToDate utilization

Gender, age and job title were significantly associated with utilization. Lower use among older lecturers is consistent with the age gradient reported among UpToDate users in other LMICs and with reviews citing age and digital health literacy as moderators of decision support adoption (Laka et al., 2021; Rosenberg et al., 2022). Differences in exposure to digital technologies, previous training, established professional routines and opportunities to use electronic resources may have contributed to the observed pattern and not necessarily less capability of using digital technologies. Younger professionals may have greater exposure to digital technologies during their training and may therefore be more accustomed to using electronic information resources. Older professionals, on the other hand, may have established ways of obtaining clinical and teaching information and may require greater exposure or support before incorporating new digital resources into their routine practice.

Teaching experience provided an interesting contrast. Lecturers with 16 years or more of teaching experience had higher utilization than those with 1–5 years of experience, even though older age was associated with lower utilization. Possible explanations include greater responsibility for curriculum, supervision and assessment that generates demand for current evidence, and self-selection of long-serving lecturers who adopt resources. Because age and experience are correlated, and because the experience and rank estimates are also based on small groups (n = 15 and n = 7), these results need replication. The unadjusted gender difference (d = 0.38) was borderline after adjustment (p = 0.055), suggesting that it is partly explained by age, rank and training. We cannot rule out a modest independent gender effect. The absence of association between attitude toward technology and use (r = −0.04) parallels Kenyan resident physicians who held favorable attitudes toward evidence-based medicine but reported limited practice because of time and skills barriers (Unadkat et al., 2021b). Perceived usefulness and workplace context, rather than general enthusiasm predicts use (Rosenberg et al., 2022). In UTAUT terms, our data suggest that the barrier lies in effort expectancy and facilitating conditions but not in generalized acceptance (Wang et al., 2025b).

### Institutional factors contributing to UpToDate utilization

The findings show that institutional factors/ support was generally positively associated with UpToDate utilization in the bivariate analysis. These findings fit evidence that a supportive professional context was among the strongest facilitators of UpToDate use, that difficulty fitting the tool into workflow was the only significant barrier(Rosenberg et al., 2022). Therefore, environment in which lecturers work may influence how visible and useful UpToDate becomes in everyday practice. Kinengyere et al. (2021) identified institutional access and active advocacy as important components of UpToDate uptake(Kinengyere et al., 2021). Rosenberg et al. (2022) similarly identified institutional and technical conditions as important facilitators or barriers to the use of digital clinical decision support tools (Rosenberg et al., 2022) .

The relationship between departmental encouragement and utilization is particularly important. Regular discussions, demonstrations and encouragement within departments may make lecturers more aware of UpToDate and provide opportunities to see how it can be used in teaching and clinical activities. Therefore, utilization may be influenced not only by the individual lecturer but also by the immediate professional environment. Evidence suggests that facilitating conditions are the strongest correlate of actual use of digital health tools by LMIC health workers(Wang et al., 2025b).

The association between integration of UpToDate into teaching and clinical follow-up and utilization provides further support for this interpretation. When a digital resource is incorporated into routine academic or clinical activities, users may have more opportunities to interact with it and may be more likely to regard it as part of their normal work. This is consistent with Unadkat et al. (2021), who suggested that perceived usefulness alone may not result in routine utilization and that integration into established workflows is important (Unadkat et al., 2021b).

Campus training was also positively associated with utilization. This finding converged with qualitative responses, where training and capacity building accounted for 21.4% of the open-ended comments. Respondents requested onsite training, mentorship, improved learning sessions and regular refresher opportunities. The convergence of the quantitative and qualitative findings suggests that training may be an important area for institutional action.

The findings concerning reliable institutional internet and work-device access require more careful consideration. Both variables were statistically significant but negatively correlated with utilization. This direction is different from what would generally be expected, since reliable connectivity and access to appropriate devices are normally considered important facilitators of digital clinical decision support use. Rosenberg et al. (2022), for example, identified poor connectivity as an important barrier(Rosenberg et al., 2022). Our exploratory analysis points to clustering. Lecturers who had not been trained rated infrastructure more favorably and reported much less departmental and campus support, so that infrastructure ratings were inversely correlated with training (r = −0.42) and with the support items (r = −0.33). The negative relationship in the current study should therefore not be interpreted to mean that reliable internet or access to work devices reduces utilization. It may reflect differences in the characteristics of respondents, measurement effects or other factors that were not captured in the study. Some campuses may have adequate connectivity and devices but have not rolled out training or promotion, so that infrastructure is necessary but clearly not sufficient. An exposure effect is also possible, in which heavier users meet and report connectivity problems that non-users never encounter.

The study further showed that although the institutional-factor composite was significantly correlated with utilization in the bivariate analysis, it was not independently associated with utilization after adjustment (b=0.091, p=0.464). This finding does not necessarily suggest that institutional support is unimportant. Rather, it may indicate that institutional and individual factors overlap in their influence on utilization. For example, institutional training and departmental encouragement may improve digital literacy and self-efficacy. Integration into teaching and clinical activities may provide opportunities for repeated use. Leadership support and institutional policies may similarly create an environment in which lecturers are encouraged to use the resource. In this way, institutional factors may influence utilization indirectly through individual skills, confidence and opportunities for use. Therefore, the findings therefore suggest that UpToDate utilization is influenced by a combination of individual and institutional circumstances.

## Implications of Findings

Findings indicated the need to move from one-off sensitization to continuous, department-based awareness activities that show specific teaching and clinical-supervision uses. Secondly, offer short, hands-on competency-based training (login and mobile access, effective searching, drug information, applying recommendations) with refreshers, targeted especially at older lecturers and those with limited exposure. Third, equip librarians and faculty champions in each department, since departmental encouragement was among the strongest correlates. Fourth, build UpToDate into assignments, case discussions and clinical follow-up so that use becomes routine and visible to students. Finally, use platform log data (registrations, log-ins by department and age) to track the effect of these measures. Because KMTC trains a large share of Kenya’s middle-level health workers, improved access among lecturers could reach many learners.

## Strengths and limitations

The high response rate and reported reliability of most scales and the convergence between the two sources of information (qualitative and quantitative), particularly regarding awareness, training and capacity building, strengthens interpretation of our findings. The study nevertheless had some limitations. Although training was associated with higher utilization, we cannot establish whether training increased utilization or whether lecturers who already used UpToDate were more likely to seek training. Second, utilization was measured as self-rated functional competence, so it may be affected by recall, social desirability and response bias. Despite these limitations, the study provides useful information for understanding of the current use of UpToDate at KMTC and for identifying areas where further awareness creation, practical training and institutional support may be useful.

## Conclusion

This study demonstrates KMTC institutional access to UpToDate but this has not translated into routine use by lecturers. Age, long teaching experience and rank are independently associated with utilization. Improving utilization may require continuous awareness building, hands-on training, departmental champions and integration of UpToDate into everyday teaching, with log-based monitoring and evaluation. Future research to focus on pragmatic trial or stepped-wedge evaluation of department-based awareness and training bundles with log-data outcomes, qualitative interviews with lecturers and librarians to understand whether lecturer use translates into students’ evidence-based practice. Taken together, sustainable approach should combine continuous awareness creation, practical skills development, mentorship, faculty engagement and meaningful integration of UpToDate into teaching and clinical activities.

## Availability of data and materials

The data used in the study is available from the corresponding author on reasonable request within existing data protection laws.

## Authors’ contributions

All authors (KM, BK, RK, GT, GA) took equal part in the study conceptualization, methodology, investigation, formal analysis, writing and review of original draft and approved the submitted version.

## Competing interests

The authors have no competing interest.

## Funding

This study received no external funding

## Data Availability

All data produced in the present study are available upon reasonable request to the authors

## Acknowledgements

We thank the lecturers who participated, the campus principals and librarians, the data collectors, and the Better Evidence Program for programme data.

## Notes

### Competing Interest Statement

The authors have declared no competing interest.

### Author Declarations

The study was approved by the KMTC Institutional Research and Ethics Review Committee and the Chuka University Ethics Committee for Research, and a research permit was issued by the National Commission for Science, Technology and Innovation (NACOSTI)

## REFERENCES

Adam, R., & MKHAI, E. (2025). The Use of Up-to-date Database by Healthcare Practitioners in Supporting Clinical Decision-Making at The Muhimbili University of Health and Allied Sciences, Tanzania. Tanzania Medical Journal, 36(1), 167–182. 10.4314/0wpabe04

Creswell JW, & Plano Clark VL. (2018). Designing and conducting mixed methods research. *3rd ed*. *Thousand Oaks*: *SAGE*; *2018*.

Gachanja, F., Mwangi, N., & Gicheru, W. (2021). E-learning in medical education during COVID-19 pandemic: experiences of a research course at Kenya Medical Training College. BMC Medical Education, 21(1), 612. 10.1186/s12909-021-03050-7

Gicheru, W., & Mwangi, N. (2023). Identifying the need to institutionalize digital equity among faculty: the experience of the Kenya Medical Training College. Frontiers in Education, 8. 10.3389/feduc.2023.1252842

Isaac, T., Zheng, J., & Jha, A. (2012). Use of *UpToDate* and outcomes in US hospitals. Journal of Hospital Medicine, 7(2), 85–90. 10.1002/jhm.944

Kinengyere, A. A., Asiimwe, G., Nyamwiza, A., Adriko, W., Twinamasiko, E., Karemani, A., & Rosenberg, J. (2025). Utilization and Uptake of the UpToDate Clinical Decision Support Tool in Five Medical Schools in Uganda (August 2022-August 2023): A Partnership with the Better Evidence Program. International Journal of Clinical Medicine, 16(02), 171–198. 10.4236/ijcm.2025.162011

Kinengyere, A. A., Rosenberg, J., Pickard, O., & Kamya, M. (2021). Utilization and uptake of the UpToDate clinical decision support tool at the Makerere University College of Health Sciences (MakCHS), Uganda. African Health Sciences, 21(2), 904–911. 10.4314/ahs.v21i2.52

KMTC. (2023). Kenya Medical Training College. (2023). *Strategic plan 2023–2028*.

Laka, M., Milazzo, A., & Merlin, T. (2021). Factors That Impact the Adoption of Clinical Decision Support Systems (CDSS) for Antibiotic Management. International Journal of Environmental Research and Public Health, 18(4), 1901. 10.3390/ijerph18041901

Rosenberg, J., Miller, K., Pickard, O., Henrich, N., Karlage, A., & Weintraub, R. (2022). Barriers and facilitators to use of a digital clinical decision support tool: a cohort study combining clickstream and survey data. BMJ Open, 12(11), e064952. 10.1136/bmjopen-2022-064952

Unadkat, M. B., Mbuba, C. K., Ngugi, A. K., & Kamya, D. (2021a). Self-reported knowledge, attitudes, practices and barriers in use of evidence-based medicine among resident physicians in Kenya: a mixed methods study. BMC Medical Education, 21(1), 537. 10.1186/s12909-021-02974-4

Unadkat, M. B., Mbuba, C. K., Ngugi, A. K., & Kamya, D. (2021b). Self-reported knowledge, attitudes, practices and barriers in use of evidence-based medicine among resident physicians in Kenya: a mixed methods study. BMC Medical Education, 21(1), 537. 10.1186/s12909-021-02974-4

Wang, M., Huang, K., Li, X., Zhao, X., Downey, L., Hassounah, S., Liu, X., Jin, Y., & Ren, M. (2025a). Health workers’ adoption of digital health technology in low- and middle-income countries: a systematic review and meta-analysis. Bulletin of the World Health Organization, 103(02), 126–135F. 10.2471/BLT.24.292157

Wolters Kluwer. (2026). UpToDate. (2026). UpToDate: Evidence-based clinical content. Wolters Kluwer. Retrieved October 5, 2026, from UpToDate.

